# Real-World Patterns of Headache Medication Use Among Service Members and Veterans With and Without Mild Traumatic Brain Injury: A LIMBIC-CENC Study

**DOI:** 10.64898/2026.11.25.26368980

**Authors:** Shirin Saleh, Erin D. Bouldin, Donald McGeary, Melissa M. Cortez, Jacob Kean, Elisabeth A. Wilde, Mary Jo Pugh

## Abstract

**Objective:** Headache is a common among service members and Veterans (SM/Vs) with mild traumatic brain injury (mTBI), yet medication use remains poorly characterized. We described headache medication patterns and examined associations with lifetime mTBI exposure.

**Setting:** The Long-Term Impact of Military-Relevant Brain Injury Consortium–Chronic Effects of Neurotrauma Consortium (LIMBIC-CENC) Prospective Longitudinal Study.

**Participants:** 1,956 SM/Vs who endorsed headaches at baseline assessment.

**Design:** Cross-sectional analysis of baseline data. Medication use was categorized as no headache-relevant medication, acute-only, preventive-only, or combination (acute and preventive) treatments. Multinomial logistic regression was used to evaluate associations between lifetime mTBI exposure and headache treatment patterns in sequentially adjusted models. Sensitivity analyses using restricted and headache-specific medication classifications were conducted to assess the robustness of findings to potential medication misclassification.

**Main Measures:** Lifetime mTBI (0, 1, or 2+) was assessed with Ohio State University TBI Identification Method; medication use was self-reported for the previous 3 months.

**Results:** Overall, 64% of participants had 2+ lifetime mTBIs. Of the participants with medication data, 45% used no headache-relevant medication, 28% acute-only, 10% preventive-only, and 17% combination therapy. Among those with severe headache impact, 52% reported no headache-relevant medication or only OTC/first-line analgesic use. In age- and sex-adjusted models (n = 1,619), 2+ mTBIs were associated with preventive-only treatment (RRR = 1.97, 95% CI 1.09–3.57) and combination therapy (RRR = 1.58, 95% CI 1.01–2.47) relative to no medication. These associations were not significant after adjustment for clinical factors. In the fully adjusted model, treatment pattern was associated with headache impact, pain interference, age, and hypertension.

Findings were generally consistent across restrictive and headache-specific medication classifications.

**Conclusion:** Nearly half of SM/V with headache used no headache-relevant medication. Associations between 2+ mTBIs and preventive-only or combination treatment were attenuated after accounting for headache impact and clinical characteristics. Routine headache assessment and longitudinal studies of treatment needs and outcomes are warranted.

## Background

Headache is a highly prevalent and often chronic symptom following traumatic brain injury (TBI), with a pooled global prevalence of approximately 49.3%.^1^ This burden is even more pronounced in the military population (56%) than in civilian populations (45%),^1–3^ and drives disability, reduced quality of life, and health care use.^4–7^ Although headache can occur following head injuries of any severity, evidence suggests it develops more frequently after mild TBI (mTBI) than moderate or severe TBI.^8–10^ Evidence also suggests a dose–response relationship with repeated mTBIs, such that individuals with a history of multiple injuries experience more severe headaches.^11,12^

Headache disorders occurring after mTBI may or may not meet formal diagnostic criteria for posttraumatic headache (PTHA). According to the International Classification of Headache Disorders-3 (ICHD-3),^13^ headaches are classified as PTHA when onset or substantial worsening occurs within seven days of the injury or recovery of consciousness. However, in cohorts with remote injury exposure, determining whether a current headache is directly attributable to a prior TBI can become increasingly challenging. Although headaches may no longer meet formal criteria for PTHA years after injury, cumulative lifetime mTBI burden may continue to influence headache severity and, in turn, medication-seeking behavior and prescribing patterns.

Headache after mTBI can be persistent and clinically complicated to treat and may not behave like “standard” primary headache. This is particularly evident in military populations, where headaches commonly present in the context of comorbid trauma-related conditions such as PTSD, which may increase headache frequency and the risk of chronicity.^8,14,15^ Veterans who sustain combined blast and blunt head trauma are significantly more likely to develop chronic headaches than those with blast-only or blunt-only injuries, indicating that cumulative or compounded injury mechanisms may increase the risk of persistent headache.^16^

Choosing pharmacotherapies for headaches after a TBI is primarily guided by phenotypic similarity to primary headache disorders,^17–19^ and the presence of comorbid conditions.^20^ Selecting a medication that can treat both the headache and a co-occurring posttraumatic symptom is a common clinical strategy. For example, amitriptyline (a tricyclic antidepressant) is often chosen when headache co-occurs with sleep disturbance because of its sedative properties.^21^ Anti-epileptic drugs such as topiramate or valproic acid are used, as they provide both headache prophylaxis and mood/seizure stabilization.^22^

Beyond comorbidities, individual injury characteristics heavily influence recovery trajectories and clinical management. Evidence suggests that individuals who have sustained a TBI demonstrate high rates of overall medication utilization, including prescribed psychotropic and pain medications.^23–26^ The number of TBIs a person sustains over their lifetime is a critical factor that impact recovery.^27^ Yet, despite the known compounding effects of repetitive trauma, little is currently known about whether cumulative lifetime mTBI exposure is associated with distinct, real-world patterns of headache medication use. Understanding treatment patterns in a real-world setting, rather than in trials where patterns are predetermined, is an important step toward understanding how headache is currently managed in this population.

This study aims to address gaps in knowledge about real-world headache treatment patterns among SM/Vs with headache. Specifically, it is unclear whether the number of lifetime mTBI is associated with particular medication-use patterns, including preventive-only or combination treatment. We hypothesized that a greater number of lifetime mTBIs would be associated with a higher likelihood of preventive-only or combination medication use, potentially reflecting more persistent or difficult-to-treat headaches. Identifying such an association could help characterize a subgroup with more complex headache management needs.

## Methods

### Study Design

This study employed an observational, cross-sectional design. All measures, including medication use, headache impact, and clinical symptoms, were collected at the baseline visit; analyses therefore describe contemporaneous associations and cannot establish whether medication patterns preceded or followed current symptoms. Data were drawn from the Long-Term Impact of Military-Relevant Brain Injury Consortium–Chronic Effects of Neurotrauma Consortium (LIMBIC-CENC), a large, geographically diverse cohort of SM/Vs with a history of combat deployment recruited between 2015 and 2024.^28^ For participants who received Veterans Health Administration care, LIMBIC-CENC data were linked to VA electronic health records through the VA Corporate Data Warehouse via the VA Informatics and Computing Infrastructure (VINCI). These records provided International Classification of Diseases (ICD) headache diagnoses, which were used in a sensitivity analysis. Detailed information on the LIMBIC-CENC study design and eligibility criteria has been published elsewhere.^28,29^ The LIMBIC-CENC study was approved by the institutional review board at each participating site, and all participants provided written informed consent before any study procedures. The University of Utah Institutional Review Board determined this secondary analysis to be exempt and waived the requirement for informed consent.

### Participants

LIMBIC-CENC eligibility required participants to be ≥18 years of age with a history of combat exposure. The parent study excluded individuals with a history of moderate-to-severe TBI or major neurological or psychiatric conditions that significantly impaired functioning (e.g., stroke, spinal cord injury, multiple sclerosis, schizophrenia, bipolar disorder), whereas common mental health conditions such as depression and PTSD were not exclusionary.^28^

Baseline data from 3,007 LIMBIC-CENC participants were initially considered for this study. After excluding 36 participants with a missing response to the headache screening item and 1,015 who did not report headache, the analytic sample included 1,956 SM/Vs who reported headache at baseline. Of these, 1,884 had medication data, and 1,619 had complete data for all model covariates and were included in the regression analyses (**Figure 1**).

**Figure 1.**
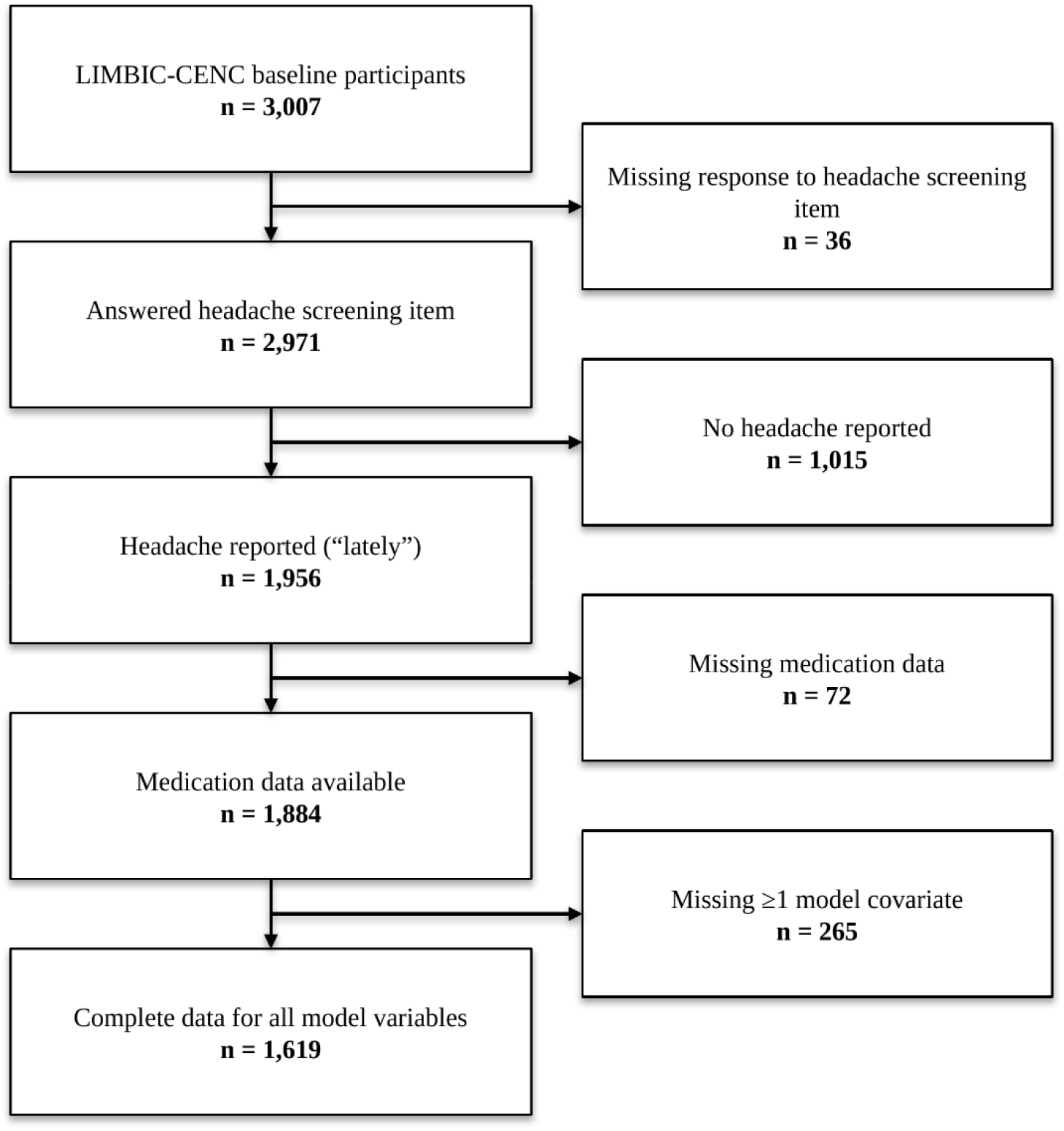
Study Sample Inclusion Flow Diagram

## Measures

### Lifetime mild TBI history (primary predictor)

Every potential concussive event (PCE) across each participant’s lifetime was identified and cataloged using a modified version of the validated Ohio State University TBI Identification (OSU TBI-ID).^30^ For the present study, cumulative lifetime mTBI exposure was classified into three mutually exclusive categories: 0 lifetime mTBIs, 1 lifetime mTBI, and ≥2 lifetime mTBIs. LIMBIC-CENC participants have heterogeneous lifetime mTBI histories, ranging from no prior mTBI to more than 10 prior injuries.

### Headache-relevant medication use (primary outcome)

Headache medication use was assessed using baseline self-reported medication data, including all prescription and over-the-counter (OTC) medications used during the previous 3 months, medication names (brand and generic), and corresponding therapeutic classes. Since the LIMBIC-CENC dataset does not specify the clinical indication for each medication, we identified medications with an established acute or preventive role in headache management (hereafter, headache-relevant medications), using a clinically informed classification scheme based on established headache guidelines and prior literature, including the American Academy of Neurology migraine prevention guidelines,^31^ the VA/DoD Clinical Practice Guideline for the Management of Headache,^32^ and the Cleveland Clinic Headache Intake Questionnaire.^33^ Medication classification was further reviewed with clinical experts (DM, MC). Both generic medication names and therapeutic class information were used to classify medications as acute or preventative.

### Headache History

For each reported TBI event, headache onset characteristics were recorded as immediate (within one week of injury) or delayed (more than one week after injury. Because participants could report multiple lifetime TBI events, they could have both immediate and delayed headache onset across different injuries. However, these self-reported onset characteristics were insufficient to establish a diagnosis of posttraumatic headache according to ICHD-3 criteria. Formal headache diagnoses and information on headache frequency, duration, and chronicity were not collected.

### Headache Impact Test-6 (HIT-6).^34^

The HIT-6 included a stem question (“Lately, have you experienced headaches?”) to identify participants reporting recent headaches, which was used to define the analytic sample for headache-related analyses. No specific timeframe was defined for the term “lately”; participants interpreted this based on their own judgment. HIT-6 symptom burden uses a 6-item self-report measure assessing pain, functional limitations, and emotional consequences. Scores range from 36 to 78, with higher scores indicating greater headache impact. Established categories include little or no impact (≤49), some impact (50–55), substantial impact (56–59), and severe impact (60–78).

### Self-reported Behavioral Symptoms

The Posttraumatic Stress Disorder Checklist (PCL-5)^35^ is a 20-item self-report measure of PTSD symptoms across four DSM-5 clusters: intrusion, avoidance, negative alteration in cognition and mood, and alterations in arousal and reactivity. Consistent with established recommendations, a total score of 33 or higher was used to identify participants with probable PTSD.^36^ The Patient Health Questionnaire (PHQ-9) assessed self-reported depression symptoms over the past 2 weeks.^37^ Scores range from 0 (none) to 27 (severe), and scores of 10 or higher indicate probable depression.^38^ Anxiety and pain interference were assessed using the corresponding Traumatic Brain Injury–Quality of Life (TBI-QOL) short-form measures, which evaluate symptoms experienced by individuals with brain injury.^39^ Raw scores were summed and converted to standardized T-scores (mean = 50, SD = 10) using the established TBI-QOL scoring procedures.^40,41^ For these subscales, a T-score of 50 represents the average score of the general population, with higher T-scores indicating greater anxiety symptoms or pain interference. TBI-QOL T-scores were analyzed as continuous variables to preserve the full range of information. Subjective sleep quality over the past month was evaluated with the Pittsburgh Sleep Quality Index (PSQI). Scores range from 0 to 21, with higher scores indicating poorer sleep quality; a global score >5 indicates poor sleep quality.^42^

### Self-reported Physical Symptoms

Using items from the Behavioral Risk Factor Surveillance System (BRFSS) questionnaire,^43^ participants reported the presence of hypertension and musculoskeletal conditions, which were analyzed as binary variables (yes/no).

### Demographics

Each participant completed a standard questionnaire to obtain information, including age, sex, race, ethnicity, and education.

### Military Characteristics and Exposures

Military characteristics included service branch, military component, and VA service-connected disability rating. Military exposure variables included history of combat-related mTBI, history of blast-related mTBI, and years since the most recent mTBI. These variables were obtained from the LIMBIC-CENC baseline survey and TBI event-level dataset.

## Statistical Analysis

Descriptive analyses were conducted in the headache-restricted cohort to characterize participant sociodemographic, military, and clinical characteristics across mTBI exposure groups (0 exposure, 1 mTBI, and 2+ mTBIs). Characteristics were summarized as means and standard deviations (SDs) for continuous variables and frequencies and percentages for categorical variables. Differences across mTBI exposure groups were assessed using one-way analysis of variance for continuous variables and chi-square tests for categorical variables.

After medication classification, the prevalence of each headache medication class was calculated as the proportion of participants with headache who reported using at least one medication within that class. Benzodiazepines and opioid/butalbital-containing products were identified but were not used to define treatment groups, and medications used predominantly for non-headache indications (e.g., gabapentinoids, SSRIs, duloxetine, mirtazapine, muscle relaxants, and corticosteroids) were not classified as headache-relevant. To examine overall treatment patterns, participants were categorized into one of four mutually exclusive treatment groups: no headache-relevant medication, acute-only (≥1 acute, no preventive), preventive-only (≥1 preventive, no acute), and combination therapy (≥1 acute and ≥1 preventive). These treatment patterns served as the primary outcome of the study, and participant characteristics were compared across treatment groups.

To describe medication use across the enrollment period, we calculated the prevalence of each headache-relevant medication class by baseline enrollment year among participants with headache and medication data (N = 1,884), using the number of participants enrolled in each year as the denominator. Each participant contributed baseline data only, so these descriptive analyses compare enrollment cohorts rather than changes within participants.

We used multivariable multinomial logistic regression to examine the association between cumulative lifetime mTBI exposure and headache treatment pattern. In addition to bivariate analysis, we fit three multivariable models in a nested fashion. In Model 1, we adjusted for age and sex. Model 2 additionally adjusted for hypertension, PTSD, depression, anxiety, sleep quality, and headache impact. Model 3 further adjusted for the pain interference to account for pain-related functional limitations potentially associated with headache and other pain conditions. Covariates were selected a priori based on prior literature and a conceptual diagram (Supplementary **Figure S1**). Because several clinical characteristics, including PTSD, depression, anxiety, sleep quality, headache impact, and pain interference, may lie along pathways linking mTBI exposure to medication use, Models 2 and 3 were interpreted as estimating associations conditional on these measured characteristics rather than the overall association between lifetime mTBI exposure and medication patterns.

Results are presented as relative risk ratios (RRRs) with 95% confidence intervals (CIs) for 1 mTBI and 2+ mTBIs, compared with 0 lifetime mTBI as the reference category. Age, anxiety and pain interference were modeled as continuous variables to retain information, whereas PTSD, depression, sleep quality, and headache impact were modeled using established clinical cutoffs to aid clinical interpretation.

The RRRs represent the relative change in the likelihood of belonging to each treatment category versus no headache-relevant medication, for each level of a categorical predictor compared with its reference group, or per one-unit increase in a continuous predictor. An RRR > 1 indicates an increased likelihood, while an RRR < 1 indicates a decreased likelihood. Statistical significance was determined using a *p*-value threshold of 0.05.

Because medication indications were unavailable, we conducted three sensitivity analyses to assess the robustness of our findings. First, we applied a restrictive medication classification that excluded selected medications with broad non-headache indications or limited evidence for headache prevention [e.g., NSAIDs, acetaminophen, antidepressants, antiseizure drugs, beta-blockers, and ACE inhibitors/ARBs]. Second, we restricted the outcome to use of relatively headache-specific treatments, including triptans, CGRP-targeting therapies, gepants/ditans, and ergot derivatives. For this analysis, we report age- and sex-adjusted estimates (Model 1) to examine associations with lifetime mTBI exposure while minimizing adjustment for clinical characteristics potentially downstream of mTBI. Third, among participants with linked VA health records (n = 1,150), we restricted the analysis to those with a documented headache diagnosis (n = 895), including migraine, tension-type headache, posttraumatic headache, or unspecified headache, and refitted Models 1 and 3 using the primary medication classification.

In a sensitivity analysis, we added baseline enrollment period (2015, 2016, 2017, 2018–2020, 2021, 2022, 2023, 2024) to Model 3 to account for changes in medication availability over time, including the introduction of CGRP-targeting therapies. 2019 and 2020 were combined with 2018 because few participants enrolled in those years.

Complete-case analysis was used for the adjusted multinomial logistic regression models. To assess potential selection bias, characteristics of participants included in and excluded from the adjusted models were compared using t-tests for continuous variables and chi-square tests for categorical variables. All analyses were conducted using R version 4.4.2.

## Results

### Sample

A total of 1,956 SM/Vs with headache were included in the analytic cohort. Lifetime mTBI exposure was available for all participants. Of these, 264 (13%) had no lifetime mTBI exposure, 443 (23%) had experienced one mTBI, and 1,249 (64%) had experienced 2+ mTBIs. Among participants with 2+ mTBIs, the median number of lifetime mTBIs was 3 (IQR 2–4; range 2–17); 38% had two, 43% had three or four, and 19% had five or more. Most participants were male (85%), and the mean age was 43 years (SD = 10). Participants with +2 mTBIs had a shorter time since their most recent injury than those with one mTBI (mean 11 vs. 16 years, *p* < 0.001). Combat-related TBI was reported by 61% of the participants, and 43% reported a history of blast exposure.

Anxiety and pain interference differed across the lifetime mTBI exposure groups (both *p*< 0.001), with lower scores among participants without mTBI (anxiety 53 vs. 57 in both mTBI groups; pain interference 56 vs. 59 and 60 for 1 and 2+ mTBIs). Sleep quality and probable PTSD also differed across groups: poor sleep (92% vs. 82%; p < 0.001) and probable PTSD (46% vs. 35%; p = 0.004) were more common among participants with 2+ mTBIs than among those without mTBI.

Among participants with a lifetime mTBI, headache onset was most often immediate only (63% in both the 1 and 2+ mTBI groups). Overall, 51% of participants reported severe headache impact, with the highest prevalence observed among those with 2+ mTBIs (53%) (**Table 1**).

**Table 1.**
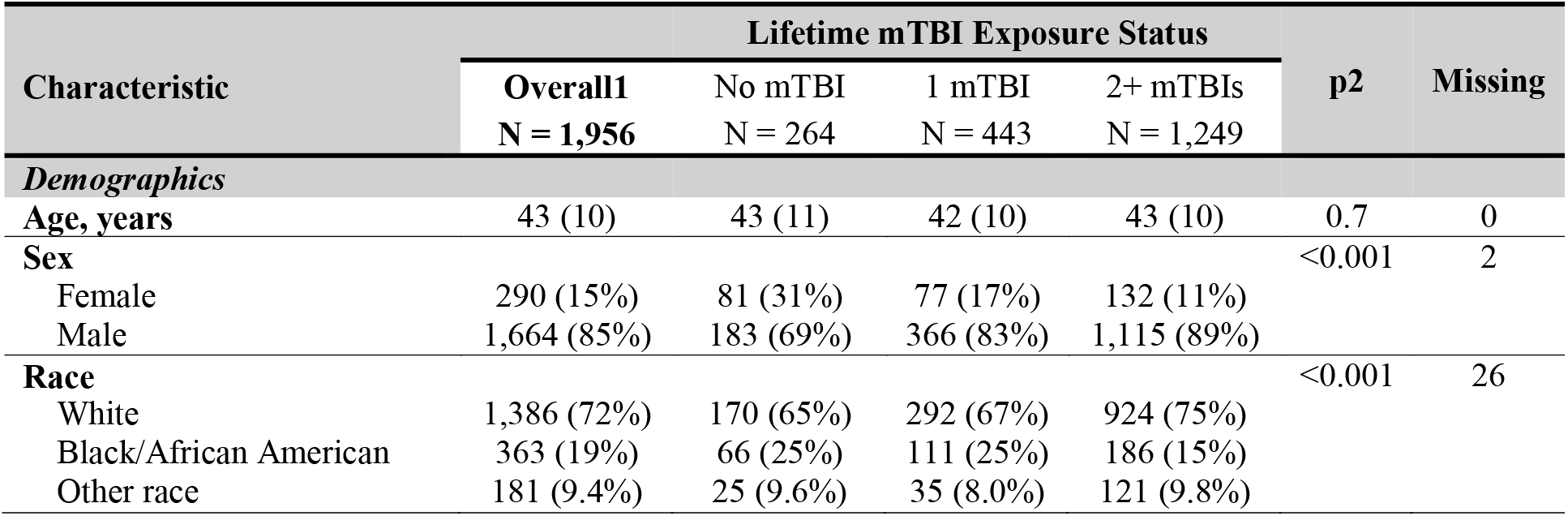
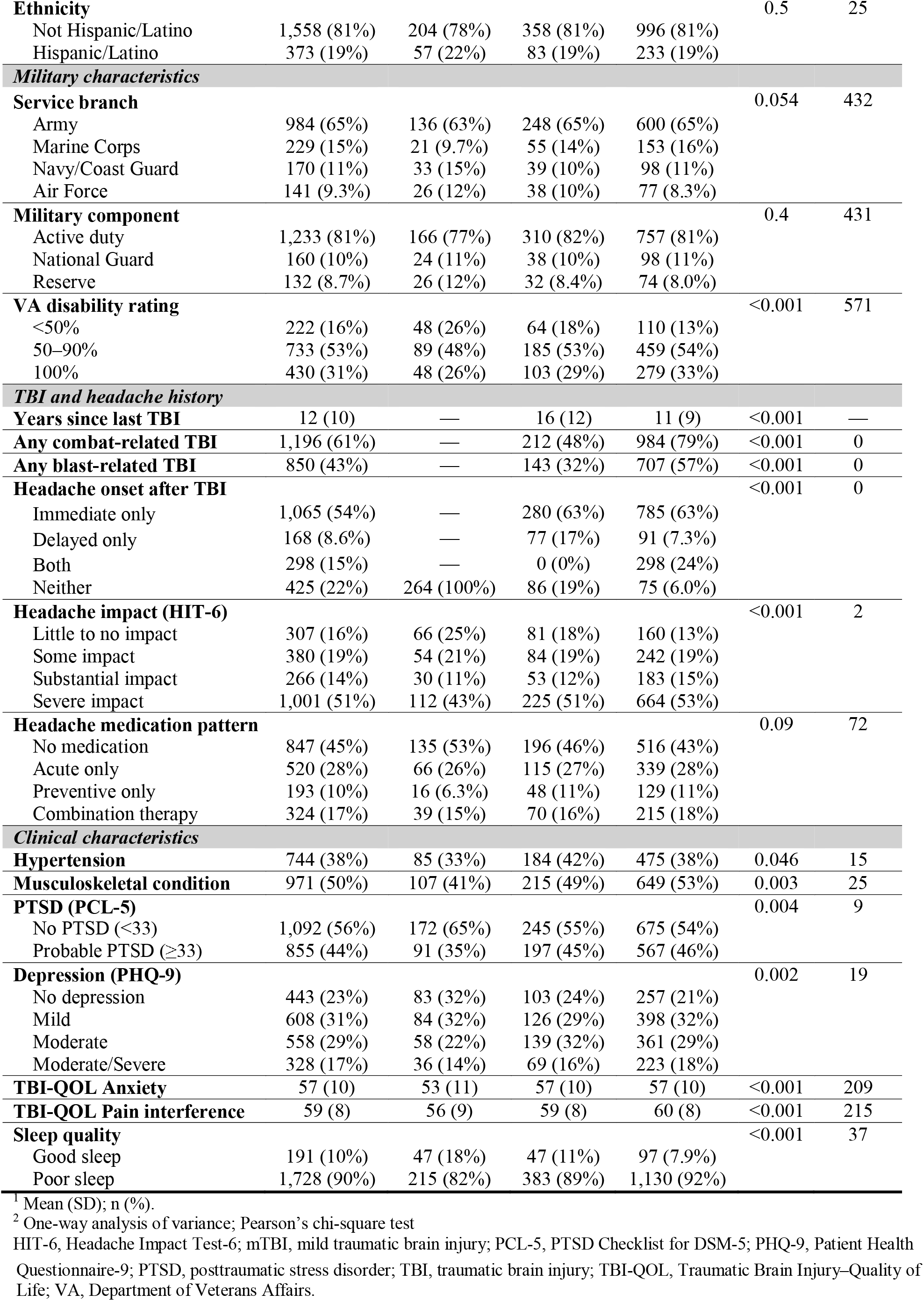
Characteristics of participants with headache by lifetime mTBI exposure.

| **Characteristic** |  | **Lifetime mTBI Exposure Status** |  |  | **p2** | **Missing** |
|---|---|---|---|---|---|---|
|  | **Overall1**<br>**N = 1,956** | No mTBI<br>N = 264 | 1 mTBI<br>N = 443 | 2+ mTBIs<br>N = 1,249 |  |  |
| *Demographics* |  |  |  |  |  |  |
| **Age, years** | 43 (10) | 43 (11) | 42 (10) | 43 (10) | 0.7 | 0 |
| **Sex** |  |  |  |  | <0.001 | 2 |
| Female | 290 (15%) | 81 (31%) | 77 (17%) | 132 (11%) |  |  |
| Male | 1,664 (85%) | 183 (69%) | 366 (83%) | 1,115 (89%) |  |  |
| **Race** |  |  |  |  | <0.001 | 26 |
| White | 1,386 (72%) | 170 (65%) | 292 (67%) | 924 (75%) |  |  |
| Black/African American | 363 (19%) | 66 (25%) | 111 (25%) | 186 (15%) |  |  |
| Other race | 181 (9.4%) | 25 (9.6%) | 35 (8.0%) | 121 (9.8%) |  |  |
| **Ethnicity** |  |  |  |  | 0.5 | 25 |
| Not Hispanic/Latino | 1,558 (81%) | 204 (78%) | 358 (81%) | 996 (81%) |  |  |
| Hispanic/Latino | 373 (19%) | 57 (22%) | 83 (19%) | 233 (19%) |  |  |
| *Military characteristics* |  |  |  |  |  |  |
| **Service branch** |  |  |  |  | 0.054 | 432 |
| Army | 984 (65%) | 136 (63%) | 248 (65%) | 600 (65%) |  |  |
| Marine Corps | 229 (15%) | 21 (9.7%) | 55 (14%) | 153 (16%) |  |  |
| Navy/Coast Guard | 170 (11%) | 33 (15%) | 39 (10%) | 98 (11%) |  |  |
| Air Force | 141 (9.3%) | 26 (12%) | 38 (10%) | 77 (8.3%) |  |  |
| **Military component** |  |  |  |  | 0.4 | 431 |
| Active duty | 1,233 (81%) | 166 (77%) | 310 (82%) | 757 (81%) |  |  |
| National Guard | 160 (10%) | 24 (11%) | 38 (10%) | 98 (11%) |  |  |
| Reserve | 132 (8.7%) | 26 (12%) | 32 (8.4%) | 74 (8.0%) |  |  |
| **VA disability rating** |  |  |  |  | <0.001 | 571 |
| <50% | 222 (16%) | 48 (26%) | 64 (18%) | 110 (13%) |  |  |
| 50–90% | 733 (53%) | 89 (48%) | 185 (53%) | 459 (54%) |  |  |
| 100% | 430 (31%) | 48 (26%) | 103 (29%) | 279 (33%) |  |  |
| *TBI and headache history* |  |  |  |  |  |  |
| **Years since last TBI** | 12 (10) | — | 16 (12) | 11 (9) | <0.001 | — |
| **Any combat-related TBI** | 1,196 (61%) | — | 212 (48%) | 984 (79%) | <0.001 | 0 |
| **Any blast-related TBI** | 850 (43%) | — | 143 (32%) | 707 (57%) | <0.001 | 0 |
| **Headache onset after TBI** |  |  |  |  | <0.001 | 0 |
| Immediate only | 1,065 (54%) | — | 280 (63%) | 785 (63%) |  |  |
| Delayed only | 168 (8.6%) | — | 77 (17%) | 91 (7.3%) |  |  |
| Both | 298 (15%) | — | 0 (0%) | 298 (24%) |  |  |
| Neither | 425 (22%) | 264 (100%) | 86 (19%) | 75 (6.0%) |  |  |
| **Headache impact (HIT-6)** |  |  |  |  | <0.001 | 2 |
| Little to no impact | 307 (16%) | 66 (25%) | 81 (18%) | 160 (13%) |  |  |
| Some impact | 380 (19%) | 54 (21%) | 84 (19%) | 242 (19%) |  |  |
| Substantial impact | 266 (14%) | 30 (11%) | 53 (12%) | 183 (15%) |  |  |
| Severe impact | 1,001 (51%) | 112 (43%) | 225 (51%) | 664 (53%) |  |  |
| **Headache medication pattern** |  |  |  |  | 0.09 | 72 |
| No medication | 847 (45%) | 135 (53%) | 196 (46%) | 516 (43%) |  |  |
| Acute only | 520 (28%) | 66 (26%) | 115 (27%) | 339 (28%) |  |  |
| Preventive only | 193 (10%) | 16 (6.3%) | 48 (11%) | 129 (11%) |  |  |
| Combination therapy | 324 (17%) | 39 (15%) | 70 (16%) | 215 (18%) |  |  |
| *Clinical characteristics* |  |  |  |  |  |  |
| **Hypertension** | 744 (38%) | 85 (33%) | 184 (42%) | 475 (38%) | 0.046 | 15 |
| **Musculoskeletal condition** | 971 (50%) | 107 (41%) | 215 (49%) | 649 (53%) | 0.003 | 25 |
| **PTSD (PCL-5)** |  |  |  |  | 0.004 | 9 |
| No PTSD (<33) | 1,092 (56%) | 172 (65%) | 245 (55%) | 675 (54%) |  |  |
| Probable PTSD (≥33) | 855 (44%) | 91 (35%) | 197 (45%) | 567 (46%) |  |  |
| **Depression (PHQ-9)** |  |  |  |  | 0.002 | 19 |
| No depression | 443 (23%) | 83 (32%) | 103 (24%) | 257 (21%) |  |  |
| Mild | 608 (31%) | 84 (32%) | 126 (29%) | 398 (32%) |  |  |
| Moderate | 558 (29%) | 58 (22%) | 139 (32%) | 361 (29%) |  |  |
| Moderate/Severe | 328 (17%) | 36 (14%) | 69 (16%) | 223 (18%) |  |  |
| **TBI-QOL Anxiety** | 57 (10) | 53 (11) | 57 (10) | 57 (10) | <0.001 | 209 |
| **TBI-QOL Pain interference** | 59 (8) | 56 (9) | 59 (8) | 60 (8) | <0.001 | 215 |
| **Sleep quality** |  |  |  |  | <0.001 | 37 |
| Good sleep | 191 (10%) | 47 (18%) | 47 (11%) | 97 (7.9%) |  |  |
| Poor sleep | 1,728 (90%) | 215 (82%) | 383 (89%) | 1,130 (92%) |  |  |

<sup>1</sup> Mean (SD); n (%).
<sup>2</sup> One-way analysis of variance; Pearson’s chi-square test
HIT-6, Headache Impact Test-6; mTBI, mild traumatic brain injury; PCL-5, PTSD Checklist for DSM-5; PHQ-9, Patient Health Questionnaire-9; PTSD, posttraumatic stress disorder; TBI, traumatic brain injury; TBI-QOL, Traumatic Brain Injury–Quality of Life; VA, Department of Veterans Affairs.

Of the 1,956 participants, 1,619 (83%) had complete data and were included in the fully adjusted model. Headache medication data were missing for 72 participants (4%). Missingness for clinical covariates was ≤2%, except for anxiety (11%) and pain interference (11%) scores. Included and excluded participants were similar in sex, ethnicity, mTBI exposure, hypertension, headache impact, and treatment pattern. Excluded participants were slightly older (mean 44 vs. 42 years; p = 0.035), had higher anxiety T-scores (59 vs. 57; p = 0.006), and were more likely to have probable PTSD (51% vs. 42%; p = 0.004) (Supplementary **Table S1**).

### Patterns of Headache Medication

Among 1,884 participants with medication data, 847 (45%) reported no headache-relevant medication use, 520 (28%) acute-only treatment, 193 (10%) preventive-only treatment, and 324 (17%) combination therapy. Participants could be counted in more than one medication class. NSAIDs were the most commonly used medication class (35%), followed by triptans (12%) and acetaminophen (10%). Among medications with preventive indications, ACE inhibitors/ARBs (7.6%), beta-blockers (7.1%), antidepressants (7.0%), and antiseizure medications (6.6%) were used at similar rates, and headache-relevant supplements were reported by 4.8% of participants.

Nearly all ACE inhibitor/ARB use was lisinopril (141 of 144 participants), and 16 of 134 beta-blocker users took atenolol or nadolol; because these agents have limited evidence for headache prevention, they were excluded from the restrictive classification, as were supplements (Supplementary **Table S2**). Only 37 of 1,937 participants (1.9%) reported a seizure disorder, suggesting that seizure treatment was unlikely to account for most antiseizure medication use in this cohort. Migraine-specific therapies other than triptans were uncommon, including onabotulinumtoxinA (1.2%), CGRP monoclonal antibodies (0.7%), gepants/ditans (0.4%), and ergot derivatives (< 0.1%); antiemetics were reported by 1.2% (**Figure 2**). Opioid/butalbital-containing medications (8%) and benzodiazepines (4%) were recorded descriptively but did not contribute to treatment group definitions.

**Figure 2.**
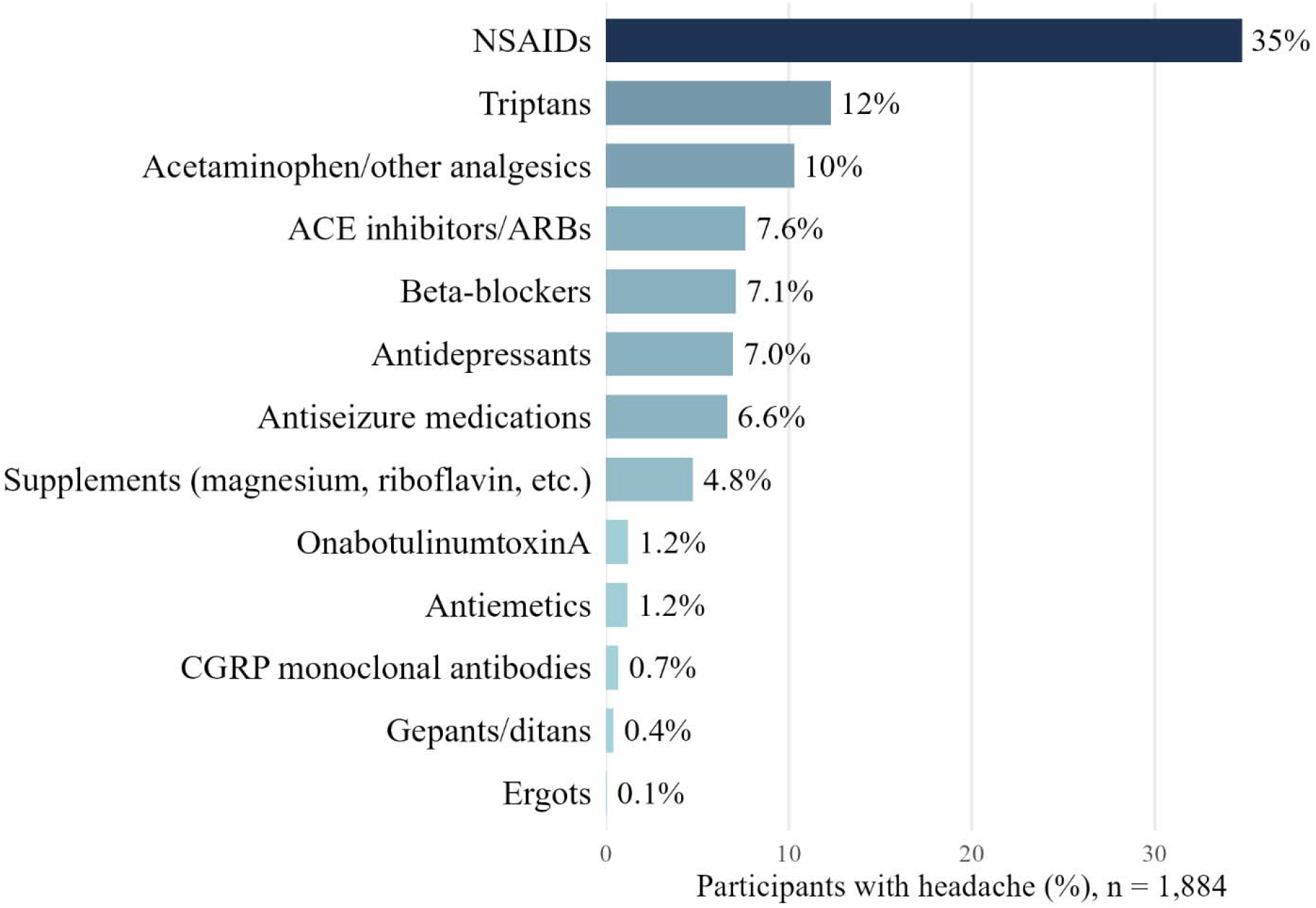
Prevalence of headache-relevant medication classes among participants with headache. A participant can appear in more than one class.

Two participants with medication data had no HIT-6 score, so the following analyses include 1,882 participants. Among participants with severe headache impact (n = 957), 338 (35%) reported no headache-relevant medication use, compared with 60% of those with little or no impact (p < 0.001), and combination therapy was more common in this group (24% vs. 6%) (Supplementary **Table S3**, **Panel A**). Overall, 494 of 957 participants with severe headache impact (52%) reported no headache-relevant medication or used only OTC/first-line analgesics. Severe headache impact was also common across all treatment patterns, reported by 40% of participants with no headache-relevant medication, 53% with acute-only, 61% with preventive-only, and 70% with combination therapy (Supplementary **Table S3, Panel B**). Among participants using any headache-relevant medication (n = 1,035), use of NSAIDs and/or acetaminophen only decreased with greater headache impact, from 53% of those with little or no impact to 25% of those with severe impact (p < 0.001) (Supplementary **Table S3, Panel C**).

Headache treatment patterns did not differ significantly by lifetime mTBI exposure (*p* = 0.09; **Table 1**). Compared with participants without mTBI, those with 2+ mTBIs had a numerically lower rate of no headache-relevant medication (43% vs. 53%) and slightly higher rates of preventive-only treatment (11% vs. 6%) and combination therapy (18% vs. 15%). Use of individual medication classes also did not differ significantly by lifetime mTBI exposure (all p ≥ 0.09; Supplementary **Table S4**). The largest numerical differences between participants without mTBI and those with 2+ mTBIs were for antiseizure medications (3.9% vs. 7.1%), opioid/butalbital-containing medications (5.5% vs. 8.4%), and botulinum toxin (0% vs. 1.6%), although use of antiseizure medications (3.9% vs. 7.1%), opioid/butalbital-containing medications (5.5% vs. 8.4%), and botulinum toxin (0% vs. 1.6%) was numerically higher among participants with 2+ mTBIs than among those without mTBI.

We also compared headache-relevant medication use across baseline enrollment years from 2015 to 2024 (Supplementary **Figure S2**). Few participants were enrolled in 2019 (n = 10) and 2020 (n = 8), so these years were combined with 2018. NSAIDs remained the most commonly used class in every period (28% to 43%), with no consistent trend over time. Use of triptans (10%–17%), ACE inhibitors/ARBs (6%–10%), beta-blockers (6%–10%), and antidepressants (5%–8%) was relatively stable. In contrast, use of opioid/butalbital-containing medications declined from 19% in 2015 to about 5% or less from 2021 onward, reaching 1.3% in 2024. Antiseizure medication use also declined, from 11% in 2015 to 2%–3% in 2023–2024, as did acetaminophen use, from 12%–14% in 2015–2016 to 6% in 2024. CGRP monoclonal antibody/atogepant use first appeared in 2021 and remained low (0.4%–2.6% per year), consistent with their recent introduction into clinical practice;.

### Multinomial Logistic Regression

Results of the sequential models, all fitted in the same complete-case sample (n = 1,619), are shown in **Table 2**. In unadjusted analyses, compared with participants without lifetime mTBI, those with two or more mTBIs had a higher relative risk of preventive-only treatment versus no headache-relevant medication (RRR = 1.85, 95% CI 1.04–3.30; p = 0.037); no other mTBI associations were significant. After adjustment for age and sex (Model 1), two or more mTBIs were associated with preventive-only treatment (RRR = 1.97, 95% CI 1.09–3.57; p = 0.024) and combination therapy (RRR = 1.58, 95% CI 1.01–2.47; p = 0.044). These associations were attenuated and no longer significant after further adjustment for hypertension, PTSD, depression, anxiety, sleep quality, and headache impact (Model 2; Supplementary **Table S5**) and for pain interference (Model 3). Compared with little or no impact, severe headache impact was associated with a higher relative risk of combination therapy (RRR = 4.33, 95% CI 2.36–7.95), and hypertension with preventive-only (RRR = 2.72, 95% CI 1.90–3.90) and combination therapy (RRR = 2.96, 95% CI 2.18–4.03). Full model estimates are provided in Supplementary **Table S5** and **S6**.

**Table 2.** Multinomial logistic regression of headache medication pattern on lifetime mTBI exposure.

| **Model** | **Lifetime mTBI (vs. none)** | **Acute only vs. none** | **Preventive only vs. none** | **Combination therapy vs. none** |
|---|---|---|---|---|
| **Unadjusted** | 1 mTBI | 1.23 (0.82–1.86) | 1.68 (0.88–3.20) | 1.23 (0.75–2.02) |
|  | 2+ mTBIs | 1.35 (0.94–1.93) | 1.85 (1.04–3.30)* | 1.46 (0.95–2.24) |
| **Model 1** | 1 mTBI | 1.26 (0.83–1.91) | 1.76 (0.92–3.38) | 1.30 (0.78–2.15) |
|  | 2+ mTBIs | 1.39 (0.96–2.01) | 1.97 (1.09–3.57)* | 1.58 (1.01–2.47)* |
| **Model 2** | 1 mTBI | 1.18 (0.77–1.80) | 1.45 (0.75–2.82) | 0.95 (0.56–1.62) |
|  | 2+ mTBIs | 1.25 (0.86–1.83) | 1.66 (0.90–3.04) | 1.13 (0.71–1.81) |
| **Model 3** | 1 mTBI | 1.14 (0.74–1.74) | 1.43 (0.73–2.78) | 0.91 (0.54–1.56) |
|  | 2+ mTBIs | 1.19 (0.81–1.74) | 1.61 (0.87–2.95) | 1.05 (0.66–1.69) |

Values are relative risk ratios (RRR) with 95% confidence intervals. \* p < 0.05.
**Model 1:** adjusted for age and sex. **Model 2:** Model 1 plus hypertension, PTSD, depression, anxiety, sleep quality, and headache impact. **Model 3:** Model 2 plus pain interference.

### Sensitivity analyses

Findings were generally consistent across alternative medication classifications and the subgroup with a documented headache diagnosis (Supplementary **Table S7**). Under the restrictive classification, 152 of 1,884 participants (8%) changed treatment group: 66 from preventive-only to no headache-relevant medication and 86 from combination to acute-only therapy. After adjustment for age and sex, 2+ mTBIs remained associated with combination therapy (RRR = 1.69, 95% CI 1.02–2.81), whereas the association with preventive-only therapy was similar in magnitude but not statistically significant (RRR = 1.82, 95% CI 0.90–3.69). These associations attenuated in the fully adjusted model, with no significant overall association between lifetime mTBI and medication patterns (p = 0.94). Use of relatively headache-specific treatments (n = 222, 14%) was not significantly associated with lifetime mTBI after adjustment for age and sex (2+ mTBIs: OR = 1.47, 95% CI 0.93–2.39). Among participants with a documented headache diagnosis in VA records (n = 757), associations were also not statistically significant, although the estimate for preventive-only treatment among those with 2+ mTBIs was elevated in Model 1 (RRR = 2.67, 95% CI 0.98–7.27).

Results were unchanged after additional adjustment for enrollment period. Lifetime mTBI remained unassociated with treatment pattern, and the mTBI RRRs were nearly identical to those in Model 3 (e.g., 2+ mTBIs, preventive only: RRR = 1.61, 95% CI 0.87–2.96).

## Discussion

This observational cross-sectional study used LIMBIC-CENC baseline data to characterize headache medication patterns among SM/Vs with headache and to examine whether lifetime mTBI exposure was associated with these patterns. Even though mTBIs were mostly remote (mean 12.2 years since the last mTBI, SD 9.8), a greater number of lifetime mTBIs was associated with more impactful headache when headache was endorsed. This finding is in line with prior studies showing a dose-response relationship between TBI severity and poor headache outcomes in SM/Vs.^11^

Notably, about one-third of participants with severe headache impact reported no headache-relevant medication, which may indicate unmet treatment need. Because medication use and headache impact were measured at the same time, this finding may also reflect effective nonpharmacologic care, prior medication intolerance or treatment failure, or personal preference. Limited access to specialty care, regional variability in specialty referrals, and costs have been linked to undertreatment of headaches in other samples.^6,44^ Stigma and health-seeking barriers have been repeatedly identified among Veterans;^45,46^ these factors may contribute for some participants, but our data cannot determine how often.

NSAIDs were the most frequently reported medication category, followed by triptans, and acetaminophen, consistent with longitudinal findings of frequent OTC and first-line analgesic use after mTBI.^47^ The low use of CGRP-targeting therapies likely reflects their recent introduction: their use in the United States rose about tenfold between 2018 and 2022, but insurers generally require two or three failed preventives first,^48^ and they were recognized as a first-line preventive option only in 2024.^49^ Evidence for their use in post-traumatic headache also remains mixed,^50,51^ which may further limit their use in people with mTBI.

Approximately half of participants with severe headache impact were either untreated or using only OTC/first-line analgesics, although use of these agents alone decreased as headache impact increased. Because dose, frequency, and duration of use were not available, we could not assess medication overuse. Nonetheless, frequent acute analgesic use among individuals with persistent, high-impact headache can raise the risk of medication-overuse headache,^7,52^ and may be worth assessing in clinical follow-up.

Combination therapy was concentrated among participants with the greatest headache burden: 70% of combination therapy users reported severe headache impact, and severe impact was associated with a fourfold higher relative risk of combination therapy in the fully adjusted model. Combined acute and preventive treatment is consistent with guideline-recommended care for frequent or disabling headache,^53^ particularly when first-line treatments are insufficient. However, concerns may arise when individuals rely on multiple acute or over-the-counter analgesics alongside preventive medications without clear optimization of therapy.^6,52^

Finally, after adjustment for age and sex, 2+ mTBIs were associated with preventive-only treatment and combination therapy, but these associations were attenuated and no longer significant after further adjustment for clinical characteristics and headache impact. Because all models used the same sample, this attenuation reflects the added covariates. Headache impact increased with mTBI number and was strongly associated with treatment, so the attenuation is consistent with mTBI influencing treatment partly through headache burden. However, psychiatric symptoms and sleep problems may also precede injury, and a cross-sectional design cannot separate mediation from confounding or overadjustment (Supplementary **Figure S1**). The fully adjusted results should therefore not be read as showing that mTBI does not influence treatment.

Sensitivity analyses findings suggest that the overall interpretation was relatively robust to alternative medication definitions and restriction to participants with documented headache diagnoses, although uncertainty regarding prescribing indications and headache phenotypes remains. Estimates for preventive-only treatment remained elevated and imprecise in the diagnosis-restricted subgroup. In the fully adjusted primary model, medication patterns were associated with older age, hypertension, greater headache impact, and higher pain interference. The association with hypertension warrants caution because some antihypertensives, including beta-blockers and ACE inhibitors/ARBs, may be prescribed for conditions other than headache. A previous VHA study reported increasing use of preventive migraine medications over time after adjustment for demographic and clinical characteristics.^54^ Although that study did not examine mTBI history, its findings further highlight the importance of considering factors beyond injury exposure when interpreting medication patterns.

Pharmacologic management of headache is highly individualized and should account for headache characteristics, symptom burden, comorbidities, contraindications, and patient preferences.^32^ Given the high burden of pain, psychiatric comorbidities, and central nervous system polypharmacy among Veterans with mTBI,^55,56^ headache medication patterns are best interpreted in the context of overall clinical burden rather than mTBI history alone.

## Limitations

This study has several limitations. First, medication indications were unavailable, and medications classified as headache-relevant may have been used for other conditions. Although sensitivity analyses generally supported the primary findings, they could not eliminate potential misclassification. Headache status was based on self-report rather than a clinical diagnosis, and information on headache phenotype, frequency, duration, and chronicity was unavailable. Thus, the cohort likely included both primary headache disorders and headaches potentially attributable to prior TBI, limiting generalizability to patients with confirmed posttraumatic headache. Additionally, baseline assessments spanned 2015–2024, during which newer treatments, including CGRP-targeting therapies, became available. Although their use was uncommon in this cohort, medication patterns may differ in more recent populations.

Second, the cross-sectional design limits temporal and causal interpretation. Medication use was reported for the preceding three months, whereas headache impact reflected recent symptoms, which may have been influenced by ongoing treatment. Furthermore, clinical characteristics included in adjusted models may lie along pathways linking mTBI exposure to medication use, potentially contributing to overadjustment. Consequently, the findings cannot establish whether observed patterns reflect undertreatment, treatment failure, or appropriate care. Third, self-reported medication use may be subject to recall bias and did not capture adherence, dosage, or frequency of acute medication use. Finally, the cohort’s military characteristics and high comorbidity burden may limit generalizability to civilian populations.

## Conclusion

In conclusion, 2+ lifetime mTBIs were associated with preventive-only and combination therapy after adjustment for age and sex, but these associations attenuated after accounting for clinical characteristics and headache impact. This attenuation may reflect pathways linking mTBI to treatment through headache burden and co-occurring symptoms, although causal mechanisms cannot be established. About half of participants with severe headache impact reported no headache-relevant medication or used only OTC/first-line analgesics, which suggests room to improve headache assessment and evaluation of treatment needs. Longitudinal studies with data on medication indication, dose, and response are needed to determine whether current patterns reflect undertreatment, treatment failure, or appropriate care.

## Data Availability

Data are available upon reasonable request to the authors, subject to approval by the LIMBIC-CENC consortium.

This work was supported by the U.S. Department of Defense through the U.S. Army Medical Research and Materiel Command, Congressionally Directed Medical Research Programs, Psychological Health and Traumatic Brain Injury Research Program award [E01 HT9425-23-1-1046] (PI: Donald D. McGeary, PhD). Dr. Pugh was also supported by VA Health Systems Research, Research Career Scientist Award, 1IK6HX003762.

## References

1. Teng Y, Ge J. Prevalence of Headache Following Traumatic Brain Injury: A Systematic Review and Meta-Analysis. Neuropsychiatr Dis Treat. 2025;21:2293–2306. doi:10.2147/NDT.S527127

2. Hoffman JM, Lucas S, Dikmen S, et al. Natural history of headache after traumatic brain injury. J Neurotrauma. 2011;28(9):1719–1725.

3. Stacey A, Lucas S, Dikmen S, et al. Natural History of Headache Five Years after Traumatic Brain Injury. J Neurotrauma. 2017;34(8):1558–1564. doi:10.1089/neu.2016.4721

4. Roper LS, Nightingale P, Su Z, Mitchell JL, Belli A, Sinclair AJ. Disability from posttraumatic headache is compounded by coexisting posttraumatic stress disorder. J Pain Res. 2017; 10:1991–1996. doi:10.2147/JPR.S129808

5. Dumke HA. Posttraumatic Headache and Its Impact on Return to Work After Mild Traumatic Brain Injury. Journal of Head Trauma Rehabilitation. 2017;32(2):E55–E65. doi:10.1097/HTR.0000000000000244

6. McGeary D, Swan AA, Kennedy E, et al. Chronic headaches after traumatic brain injury: Diagnostic complexity associated with increased cost. NeuroRehabilitation. 2024;55(3):303–317. doi:10.3233/NRE-230277

7. Mofatteh M. Examining the association between traumatic brain injury and headache. J Integr Neurosci. 2021;20(4):1079–1094. doi:10.31083/j.jin2004109

8. Theeler B, Lucas S, Riechers RG, Ruff RL. Post-traumatic headaches in civilians and military personnel: a comparative, clinical review. Headache. 2013;53(6):881–900. doi:10.1111/head.12123

9. Herrero Babiloni A, Bouferguene Y, Exposto FG, et al. The prevalence of persistent post-traumatic headache in adult civilian traumatic brain injury: a systematic review and meta-analysis on the past 14 years. Pain. 2023;164(12):2627–2641. doi:10.1097/j.pain.0000000000002949

10. Flynn S, Moore B, van der Merwe AJ, et al. Headaches in Traumatic Brain Injury: Improvement Over Time, Associations With Quality of Life, and Impact of Migraine-Type Headaches. Journal of Head Trauma Rehabilitation. 2023;38(2):E109–E117. doi:10.1097/HTR.0000000000000790

11. Coffman C, Reyes D, Hess MC, et al. Relationship Between Headache Characteristics and a Remote History of TBI in Veterans. Neurology. 2022;99(2). doi:10.1212/WNL.0000000000200518

12. Walker WC, Clark SW, Eppich K, et al. Headache among combat-exposed veterans and service members and its relation to mild traumatic brain injury history and other factors: a LIMBIC-CENC study. Front Neurol. 2023;14. doi:10.3389/fneur.2023.1242871

13. Headache Classification Committee of the International Headache Society (IHS) The International Classification of Headache Disorders, 3rd edition. Cephalalgia. 2018;38(1):1-211. doi:10.1177/0333102417738202

14. Kozminski M. Combat-related posttraumatic headache: diagnosis, mechanisms of injury, and challenges to treatment. J Am Osteopath Assoc. 2010;110(9):514–519.

15. Waung MW, Abrams GM. Combat-Related Headache and Traumatic Brain Injury. Curr Pain Headache Rep. 2012;16(6):533–538. doi:10.1007/s11916-012-0294-7

16. Nabity PS, Jaramillo CA, Resick PA, et al. Persistent posttraumatic headaches and functioning in veterans: Injury type can matter. Headache: The Journal of Head and Face Pain. 2021;61(9):1334–1341. doi:10.1111/head.14210

17. Ashina H, Eigenbrodt AK, Seifert T, et al. Post-traumatic headache attributed to traumatic brain injury: classification, clinical characteristics, and treatment. Lancet Neurol. 2021;20(6):460–469. doi:10.1016/S1474-4422(21)00094-6

18. Ashina H, Dodick DW. Post-traumatic Headache: Pharmacologic Management and Targeting CGRP Signaling. Curr Neurol Neurosci Rep. 2022;22(2):105–111. doi:10.1007/s11910-022-01175-w

19. Larsen EL, Ashina H, Iljazi A, et al. Acute and preventive pharmacological treatment of post-traumatic headache: a systematic review. J Headache Pain. 2019;20(1):98. doi:10.1186/s10194-019-1051-7

20. Baandrup L, Jensen R. Chronic post-traumatic headache - a clinical analysis in relation to the international headache classification 2nd edition. Cephalalgia. 2005;25. doi:10.1111/j.1468-2982.2004.00818.x

21. Lucas S. Headache Management in Concussion and Mild Traumatic Brain Injury. PM&R. 2011;3(10S2). doi:10.1016/j.pmrj.2011.07.016

22. Formisano R, Bivona U, Catani S, D’Ippolito M, Buzzi MG. Post-traumatic headache: facts and doubts. J Headache Pain. 2009;10(3):145-152. doi:10.1007/s10194-009-0108-4

23. Vehviläinen J, Skrifvars MB, Reinikainen M, et al. Psychotropic medication use among patients with a traumatic brain injury treated in the intensive care unit: a multi-centre observational study. Acta Neurochir (Wien). 2021;163(10):2909–2917. doi:10.1007/s00701-021-04956-3

24. Starosta AJ, Adams RS, Marwitz JH, et al. Scoping Review of Opioid Use After Traumatic Brain Injury. Journal of Head Trauma Rehabilitation. 2021;36(5):310–327. doi:10.1097/HTR.0000000000000721

25. Cosano G, Giangreco M, Ussai S, et al. Polypharmacy and the use of medications in inpatients with acquired brain injury during post-acute rehabilitation: A cross-sectional study. Brain Inj. 2016;30(3):353–362. doi:10.3109/02699052.2015.1118767

26. Yasseen B, Colantonio A, Ratcliff G. Prescription medication use in persons many years following traumatic brain injury. Brain Inj. 2008;22(10):752–757. doi:10.1080/02699050802320132

27. Molero Y, Sharp DJ, D’Onofrio BM, et al. Medication utilization in traumatic brain injury patients—insights from a population-based matched cohort study. Front Neurol. 2024;15. doi:10.3389/fneur.2024.1339290

28. Walker WC, Carne W, Franke LM, et al. The Chronic Effects of Neurotrauma Consortium (CENC) multi-centre observational study: Description of study and characteristics of early participants. Brain Inj. 2016;30(12):1469–1480. doi:10.1080/02699052.2016.1219061

29. Walker WC, Hirsch S, Carne W, et al. Chronic Effects of Neurotrauma Consortium (CENC) multicentre study interim analysis: Differences between participants with positive versus negative mild TBI histories. Brain Inj. 2018;32(9):1079–1089. doi:10.1080/02699052.2018.1479041

30. Corrigan JD, Bogner J. Initial Reliability and Validity of the Ohio State University TBI Identification Method. Journal of Head Trauma Rehabilitation. 2007;22(6):318–329. doi:10.1097/01.HTR.0000300227.67748.77

31. Silberstein SD, Holland S, Freitag F, Dodick DW, Argoff C, Ashman E. Evidence-based guideline update: Pharmacologic treatment for episodic migraine prevention in adults. Neurology. 2012;78(17):1337–1345. doi:10.1212/WNL.0b013e3182535d20

32. Sico JJ, Antonovich NM, Ballard-Hernandez J, et al. 2023 U.S. Department of Veterans Affairs and U.S. Department of Defense Clinical Practice Guideline for the Management of Headache. Ann Intern Med. 2024;177(12):1675-1694. doi:10.7326/ANNALS-24-00551

33. Tepper S, Tepper D. The Cleveland Clinic Manual of Headache Therapy. 2011. doi:10.1007/978-1-4614-0179-7

34. Kosinski M, Bayliss MS, Bjorner JB, et al. A six-item short-form survey for measuring headache impact: The HIT-6^TM^. Quality of Life Research. 2003;12(8):963–974. doi:10.1023/A:1026119331193

35. Blevins CA, Weathers FW, Davis MT, Witte TK, Domino JL. The Posttraumatic Stress Disorder Checklist for *DSM 5* (PCL 5): Development and Initial Psychometric Evaluation. J Trauma Stress. 2015;28(6):489–498. doi:10.1002/jts.22059

36. Forkus SR, Raudales AM, Rafiuddin HS, Weiss NH, Messman BA, Contractor AA. The Posttraumatic Stress Disorder (PTSD) Checklist for DSM-5: A Systematic Review of Existing Psychometric Evidence. Clin Psychol (New York). 2023;30(1):110–121. doi:10.1037/cps0000111

37. Kroenke K, Spitzer RL, Williams JBW. The PHQ-9: validity of a brief depression severity measure. J Gen Intern Med. 2001;16(9):606–613. doi:10.1046/j.1525-1497.2001.016009606.x

38. Manea L, Gilbody S, McMillan D. Optimal cut-off score for diagnosing depression with the Patient Health Questionnaire (PHQ-9): a meta-analysis. CMAJ. 2012;184(3):E191–6. doi:10.1503/cmaj.110829

39. Tulsky DS, Kisala PA, Victorson D, et al. The TBI-QOL: development and calibration of item banks to measure patient reported outcomes following traumatic brain injury. Journal of Head Trauma Rehabilitation. 2016;31(1):40–51. doi:10.1097/HTR.0000000000000131

40. Tulsky DS, Kisala PA. An Overview of the Traumatic Brain Injury–Quality of Life (TBI-QOL) Measurement System. Journal of Head Trauma Rehabilitation. 2019;34(5):281–288. doi:10.1097/HTR.0000000000000531

41. Lange RT, Brickell TA, Bailie JM, Tulsky DS, French LM. Clinical Utility and Psychometric Properties of the Traumatic Brain Injury Quality of Life Scale (TBI-QOL) in US Military Service Members. Journal of Head Trauma Rehabilitation. 2016;31(1):62–78. doi:10.1097/HTR.0000000000000149

42. Buysse DJ, Reynolds CF, Monk TH, Berman SR, Kupfer DJ. The Pittsburgh sleep quality index: A new instrument for psychiatric practice and research. Psychiatry Res. 1989;28(2):193–213. doi:10.1016/0165-1781(89)90047-4

43. Centers for Disease Control and Prevention (CDC). Behavioral Risk Factor Surveillance System Survey Questionnaire. 2022.

44. Swan AA, Akin FW, Amuan ME, et al. Disruptive Dizziness Among Post-9/11 Veterans With Deployment-Related Traumatic Brain Injury. Journal of Head Trauma Rehabilitation. 2022;37(4):199–212. doi:10.1097/HTR.0000000000000714

45. Haun JN, Nakase-Richardson R, Cotner BA, et al. Stakeholder Engagement to Identify Implementation Strategies to Overcome Barriers to Delivering Chronic Pain Treatments: A NIDILRR and VA TBI Model Systems Collaborative Project. Journal of Head Trauma Rehabilitation. 2024;39(1):E29–E40. doi:10.1097/HTR.0000000000000920

46. Nworah U, Symes L, Young A, Langford R. Afghanistan and Iraq War Veterans’ Health Care Needs and Their Underuse of Health Care Resources: Implications for Psychiatric-Mental Health Nurses. J Psychosoc Nurs Ment Health Serv. 2014;52(6):42–49. doi:10.3928/02793695-20140210-02

47. Ditommaso C, Hoffman JM, Lucas S, Dikmen S, Temkin N, Bell KR. Medication usage patterns for headache treatment after mild traumatic brain injury. Headache. 2014;54(3):511–519. doi:10.1111/HEAD.12254

48. Moura CS, Randall JR, Klarenbach S, et al. Persistence, switching, and healthcare use after initiating calcitonin gene-related peptide inhibitors: a real-world assessment. J Headache Pain. 2025;27(1):19. doi:10.1186/s10194-025-02167-0

49. Charles AC, Digre KB, Goadsby PJ, Robbins MS, Hershey A. Calcitonin gene related peptide targeting therapies are a first line option for the prevention of migraine: An American Headache Society position statement update. Headache: The Journal of Head and Face Pain. 2024;64(4):333–341. doi:10.1111/head.14692

50. Russo AF, Kaiser EA. CGRP and Migraine: Real-World Insights and Future Therapeutic Directions. Annu Rev Med. 2026;77(1):415-432. doi:10.1146/annurev-med-050224-111631

51. Saleh S, Sanford EM, McGeary DD, et al. Efficacy and outcomes of pharmacological treatments for headaches after traumatic brain injury: A systematic review. Cephalalgia. 2026;46(4). doi:10.1177/03331024261441563

52. Ashina H, Iljazi A, Al-Khazali HM, et al. Persistent post-traumatic headache attributed to mild traumatic brain injury: Deep phenotyping and treatment patterns. Cephalalgia. 2020;40(6):554–564. doi:10.1177/0333102420909865

53. Ailani J, Burch RC, Robbins MS, Board of Directors of the American Headache Society. The American Headache Society Consensus Statement: Update on integrating new migraine treatments into clinical practice. Headache. 2021;61(7):1021–1039. doi:10.1111/head.14153

54. Altalib H, Fenton B, Sico J, et al. Increase in migraine diagnoses and guideline-concordant treatment in veterans, 2004–2012. *Cephalalgia*. 2017;37(1):3-10. doi:10.1177/0333102416631959

55. Jaramillo CA, Eapen BC, McGeary CA, et al. A cohort study examining headaches among veterans of Iraq and Afghanistan wars: Associations with traumatic brain injury, PTSD, and depression. Headache. 2016;56(3):528–539. doi:10.1111/head.12726

56. Swan AA, Kennedy E, Cooper DB, et al. Comorbidity and polypharmacy impact neurobehavioral symptoms and symptom validity failure among post-9/11 veterans with mild traumatic brain injury. Front Neurol. 2023;14:1228377. doi:10.3389/fneur.2023.1228377

