## Supplemental Data 1 for "Real-World Patterns of Headache Medication Use Among Service Members and Veterans With and Without Mild Traumatic Brain Injury: A LIMBIC-CENC Study": Preprint_Pharmacological treatment patterns_supplementals.docx

**Table S1.** Characteristics of participants with headache included in and excluded from the adjusted regression models

| Characteristic | Excluded | Included | p-value^2^ |
| --- | --- | --- | --- |
|  | **N = 337^1^** | **N = 1,619^1^** |  |
| Age (years) | 44 (11) | 42 (10) | 0.035 |
| Sex |  |  | 0.4 |
| Female | 44 (13%) | 246 (15%) |  |
| Male | 291 (87%) | 1,373 (85%) |  |
| Race |  |  | 0.037 |
| Black/African American | 76 (23%) | 287 (18%) |  |
| Other race | 36 (11%) | 145 (9.1%) |  |
| White | 218 (66%) | 1,168 (73%) |  |
| Ethnicity |  |  | 0.4 |
| Hispanic/Latino | 70 (21%) | 303 (19%) |  |
| Not Hispanic/Latino | 262 (79%) | 1,296 (81%) |  |
| Lifetime mTBI Exposure Status |  |  | 0.2 |
| 0 exposure | 55 (16%) | 209 (13%) |  |
| 1 TBI | 73 (22%) | 370 (23%) |  |
| 2+ TBIs | 209 (62%) | 1,040 (64%) |  |
| Hypertension | 136 (42%) | 608 (38%) | 0.13 |
| Musculoskeletal Condition | 161 (50%) | 810 (50%) | >0.9 |
| Depression (PHQ-9) |  |  | 0.4 |
| Mild | 101 (32%) | 507 (31%) |  |
| Moderate | 89 (28%) | 469 (29%) |  |
| Moderate/Severe | 63 (20%) | 265 (16%) |  |
| No depression | 65 (20%) | 378 (23%) |  |
| TBI-QoL Anxiety | 59 (9) | 57 (10) | 0.006 |
| Sleep Quality |  |  | 0.2 |
| Good sleep | 23 (7.7%) | 168 (10%) |  |
| Poor sleep | 277 (92%) | 1,451 (90%) |  |
| TBI-QoL Pain Interference | 60 (8) | 59 (8) | 0.067 |
| PTSD (PCL-5) |  |  | 0.004 |
| No PTSD (<33) | 160 (49%) | 932 (58%) |  |
| PTSD (≥33) | 168 (51%) | 687 (42%) |  |
| Headache Onset Pattern after TBI | |  | <0.001 |
| Both immediate and delayed | 39 (12%) | 259 (16%) |  |
| Delayed only | 30 (8.9%) | 138 (8.5%) |  |
| Immediate only | 165 (49%) | 900 (56%) |  |
| Neither | 103 (31%) | 322 (20%) |  |
| Headache Impact |  |  | 0.2 |
| Little to no impact | 57 (17%) | 250 (15%) |  |
| Severe impact | 182 (54%) | 819 (51%) |  |
| Some impact | 52 (16%) | 328 (20%) |  |
| Substantial impact | 44 (13%) | 222 (14%) |  |
| Treatment Pattern |  |  | 0.4 |
| No headache-directed medication | 132 (50%) | 715 (44%) |  |
| Acute only | 66 (25%) | 454 (28%) |  |
| Preventive only | 26 (9.8%) | 167 (10%) |  |
| Combination therapy | 41 (15%) | 283 (17%) |  |

^1^ Mean (SD); n (%). ^2^ Welch Two Sample t-test; Pearson’s Chi-squared test.

mTBI, mild traumatic brain injury; TBI, traumatic brain injury; TBI-QoL, Traumatic Brain Injury Quality of Life; PHQ-9, Patient Health Questionnaire-9; PCL-5, PTSD Checklist for DSM-5; PTSD, posttraumatic stress disorder; HIT-6, Headache Impact Test-6; SD, standard deviation.

**Table S2.** Classification of headache-relevant medications (participants with headache, N = 1,956)

| Category | Role | Records, n | Participants, n | Medications | Restrictive analysis |
| --- | --- | --- | --- | --- | --- |
| NSAIDs | Acute | 773 | 655 | Aspirin, ibuprofen, naproxen, diclofenac, celecoxib, meloxicam, indomethacin, ketorolac; Excedrin | Included |
| Triptans | Acute | 241 | 232 | Sumatriptan, rizatriptan, eletriptan, zolmitriptan, naratriptan, frovatriptan | Included |
| Acetaminophen | Acute | 210 | 194 | Acetaminophen, Midrin | Included |
| Antiemetics | Acute | 22 | 22 | Metoclopramide, prochlorperazine, promethazine | Included |
| Gepants/ditans | Acute | 8 | 8 | Rimegepant, ubrogepant | Included |
| Ergot derivatives | Acute | 1 | 1 | Dihydroergotamine | Included |
| ACE inhibitors /ARBs | Preventive | 144 | 144 | Candesartan (n = 3) | Included |
|  |  |  |  | Lisinopril, incl. lisinopril/HCTZ (n = 141) | Excluded¹ |
| Beta-blockers | Preventive | 135 | 134 | Propranolol, metoprolol, timolol (n = 118) | Included |
|  |  |  |  | Atenolol, nadolol (n = 16) | Excluded¹ |
| Antidepressants | Preventive | 136 | 131 | Amitriptyline, nortriptyline, venlafaxine | Included |
| Antiseizure medications | Preventive | 129 | 125 | Topiramate, divalproex/valproic acid | Included |
| Supplements | Preventive | 96 | 90 | Magnesium, riboflavin, coenzyme Q10 | Excluded² |
| Botulinum toxin | Preventive | 23 | 23 | OnabotulinumtoxinA, incobotulinumtoxinA | Included |
| CGRP mAbs/atogepant | Preventive | 13 | 13 | Erenumab, fremanezumab, atogepant | Included |
| Opioids/butalbital | Not used to define groups | 171 | 143 | Butalbital (Fioricet), hydrocodone, oxycodone, codeine, tramadol, morphine | n/a |
| Benzodiazepines | Not used to define groups | 78 | 75 | Alprazolam, clonazepam, diazepam, lorazepam | n/a |

¹ Limited evidence for headache prevention and predominantly prescribed for hypertension.
² Low-certainty evidence and common general-health use.

ACE, angiotensin-converting enzyme; ARB, angiotensin receptor blocker; CGRP, calcitonin gene-related peptide; HCTZ, hydrochlorothiazide; NSAID, nonsteroidal anti-inflammatory drug.

**Table S3.** Headache impact (HIT-6) by headache-relevant medication pattern

**Panel A.** Medication pattern within each HIT-6 category (row %)

| HIT-6 category | No headache-relevant medication n (%) | Acute only n (%) | Preventive only n (%) | Combination therapy n (%) | Total n |
| --- | --- | --- | --- | --- | --- |
| Little to no impact | 179 (60%) | 75 (25%) | 24 (8.1%) | 19 (6.4%) | 297 |
| Some impact | 200 (54%) | 98 (26%) | 30 (8.1%) | 44 (12%) | 372 |
| Substantial impact | 130 (51%) | 71 (28%) | 21 (8.2%) | 34 (13%) | 256 |
| Severe impact | 338 (35%) | 275 (29%) | 118 (12%) | 226 (24%) | 957 |
| Total | 847 (45%) | 519 (28%) | 193 (10%) | 323 (17%) | 1,882 |

**Panel B.** HIT-6 category within each medication pattern (column %)

| HIT-6 category | No headache-relevant medication N = 847 | Acute only N = 519 | Preventive only N = 193 | Combination therapy N = 323 | Total N = 1,882 |
| --- | --- | --- | --- | --- | --- |
| Little to no impact | 179 (21%) | 75 (14%) | 24 (12%) | 19 (5.9%) | 297 (16%) |
| Some impact | 200 (24%) | 98 (19%) | 30 (16%) | 44 (14%) | 372 (20%) |
| Substantial impact | 130 (15%) | 71 (14%) | 21 (11%) | 34 (11%) | 256 (14%) |
| Severe impact | 338 (40%) | 275 (53%) | 118 (61%) | 226 (70%) | 957 (51%) |

**Panel C.** OTC/first-line-only treatment by HIT-6 category, among participants using any headache-relevant medication (n = 1,035; row %)

| HIT-6 category | OTC/first-line only n (%) | Other treatment n (%) | Total n |
| --- | --- | --- | --- |
| Little to no impact | 62 (53%) | 56 (47%) | 118 |
| Some impact | 75 (44%) | 97 (56%) | 172 |
| Substantial impact | 47 (37%) | 79 (63%) | 126 |
| Severe impact | 156 (25%) | 463 (75%) | 619 |
| Total | 340 (33%) | 695 (67%) | 1,035 |

Panels A and B show the same cross-tabulation: Panel A gives row percentages and Panel B gives column percentages. p < 0.001 for the association between HIT-6 category and medication pattern (Pearson’s chi-square test).
Panel C: p < 0.001 (Pearson’s chi-square test). OTC/first-line only was defined as use of NSAIDs and/or acetaminophen/simple analgesics only, with no other headache-relevant medication class; Other treatment includes all other headache-relevant medication patterns. HIT-6, Headache Impact Test-6; NSAID, nonsteroidal anti-inflammatory drug; OTC, over-the-counter.

**Table S4.** Use of headache-relevant medication classes by lifetime mTBI exposure

| Medication class | Role | Overall N = 1,884 | 0 exposure N = 256 | 1 mTBI N = 429 | 2+ mTBIs N = 1,199 | p-value^1^ |
| --- | --- | --- | --- | --- | --- | --- |
| NSAIDs | Acute | 655 (35%) | 81 (32%) | 145 (34%) | 429 (36%) | 0.4 |
| Triptans | Acute | 232 (12%) | 28 (11%) | 54 (13%) | 150 (13%) | 0.8 |
| Acetaminophen | Acute | 194 (10%) | 19 (7.4%) | 48 (11%) | 127 (11%) | 0.2 |
| Antiemetics | Acute | 22 (1.2%) | 2 (0.8%) | 4 (0.9%) | 16 (1.3%) | 0.7 |
| Gepants/ditans | Acute | 8 (0.4%) | 1 (0.4%) | 1 (0.2%) | 6 (0.5%) | 0.8 |
| Ergot derivatives | Acute | 1 (<0.1%) | 0 (0%) | 0 (0%) | 1 (<0.1%) | 0.8 |
| ACE inhibitors/ARBs | Preventive | 144 (7.6%) | 14 (5.5%) | 34 (7.9%) | 96 (8.0%) | 0.4 |
| Beta-blockers | Preventive | 134 (7.1%) | 20 (7.8%) | 30 (7.0%) | 84 (7.0%) | 0.9 |
| Antidepressants | Preventive | 131 (7.0%) | 16 (6.2%) | 34 (7.9%) | 81 (6.8%) | 0.6 |
| Antiseizure medications | Preventive | 125 (6.6%) | 10 (3.9%) | 30 (7.0%) | 85 (7.1%) | 0.2 |
| Supplements | Preventive | 90 (4.8%) | 13 (5.1%) | 19 (4.4%) | 58 (4.8%) | >0.9 |
| Botulinum toxin | Preventive | 23 (1.2%) | 0 (0%) | 4 (0.9%) | 19 (1.6%) | 0.092 |
| CGRP mAbs/atogepant | Preventive | 13 (0.7%) | 3 (1.2%) | 0 (0%) | 10 (0.8%) | 0.12 |
| Opioids/butalbital | Not used to define groups | 143 (7.6%) | 14 (5.5%) | 28 (6.5%) | 101 (8.4%) | 0.2 |
| Benzodiazepines | Not used to define groups | 75 (4.0%) | 13 (5.1%) | 13 (3.0%) | 49 (4.1%) | 0.4 |

^1^ Pearson’s chi-square test. Values are n (column %). ACE, angiotensin-converting enzyme; ARB, angiotensin receptor blocker; CGRP, calcitonin gene-related peptide; mAbs, monoclonal antibodies; mTBI, mild traumatic brain injury; NSAID, nonsteroidal anti-inflammatory drug.

**Table S5.** Multinomial logistic regression of headache medication pattern, partially adjusted (Model 2; n = 1,619)

| Characteristic | Acute only | | Preventive only | | Combination therapy | |
| --- | --- | --- | --- | --- | --- | --- |
|  | RRR (95% CI) | p-value | RRR (95% CI) | p-value | RRR (95% CI) | p-value |
| Lifetime mTBI |  |  |  |  |  |  |
| No mTBI | 1.00 (Ref) |  | 1.00 (Ref) |  | 1.00 (Ref) |  |
| 1 mTBI | 1.18 (0.77–1.80) | 0.5 | 1.45 (0.75–2.82) | 0.3 | 0.95 (0.56–1.62) | 0.9 |
| 2+ mTBIs | 1.25 (0.86–1.83) | 0.2 | 1.66 (0.90–3.04) | 0.10 | 1.13 (0.71–1.81) | 0.6 |
| Age, years | 1.04 (1.03–1.06) | <0.001 | 1.04 (1.02–1.05) | <0.001 | 1.05 (1.04–1.07) | <0.001 |
| Sex |  |  |  |  |  |  |
| Female | 1.00 (Ref) |  | 1.00 (Ref) |  | 1.00 (Ref) |  |
| Male | 0.84 (0.59–1.20) | 0.3 | 0.65 (0.40–1.06) | 0.085 | 0.64 (0.43–0.97) | 0.034 |
| Hypertension |  |  |  |  |  |  |
| No | 1.00 (Ref) |  | 1.00 (Ref) |  | 1.00 (Ref) |  |
| Yes | 1.12 (0.86–1.46) | 0.4 | 2.72 (1.90–3.90) | <0.001 | 2.96 (2.18–4.02) | <0.001 |
| PTSD (PCL-5) |  |  |  |  |  |  |
| No PTSD (<33) | 1.00 (Ref) |  | 1.00 (Ref) |  | 1.00 (Ref) |  |
| PTSD (≥33) | 0.76 (0.54–1.07) | 0.12 | 1.20 (0.74–1.95) | 0.5 | 0.97 (0.65–1.47) | >0.9 |
| Depression (PHQ-9) |  |  |  |  |  |  |
| No depression | 1.00 (Ref) |  | 1.00 (Ref) |  | 1.00 (Ref) |  |
| Mild | 0.98 (0.69–1.40) | >0.9 | 0.87 (0.51–1.50) | 0.6 | 0.94 (0.59–1.49) | 0.8 |
| Moderate | 1.11 (0.73–1.70) | 0.6 | 1.07 (0.58–2.00) | 0.8 | 0.89 (0.52–1.52) | 0.7 |
| Moderate/Severe | 1.11 (0.64–1.91) | 0.7 | 1.33 (0.63–2.84) | 0.5 | 1.30 (0.69–2.47) | 0.4 |
| TBI-QOL Anxiety, T-score | 1.00 (0.98–1.02) | >0.9 | 0.99 (0.96–1.01) | 0.2 | 1.00 (0.98–1.02) | >0.9 |
| Sleep quality |  |  |  |  |  |  |
| Good sleep | 1.00 (Ref) |  | 1.00 (Ref) |  | 1.00 (Ref) |  |
| Poor sleep | 1.42 (0.93–2.18) | 0.11 | 1.92 (0.94–3.92) | 0.074 | 1.93 (1.03–3.63) | 0.042 |
| Headache impact (HIT-6) |  |  |  |  |  |  |
| Little to no impact | 1.00 (Ref) |  | 1.00 (Ref) |  | 1.00 (Ref) |  |
| Some impact | 1.16 (0.77–1.75) | 0.5 | 1.13 (0.61–2.10) | 0.7 | 1.87 (0.99–3.52) | 0.054 |
| Substantial impact | 1.40 (0.89–2.21) | 0.14 | 1.07 (0.52–2.17) | 0.9 | 2.32 (1.17–4.58) | 0.016 |
| Severe impact | 2.15 (1.45–3.19) | <0.001 | 2.06 (1.14–3.71) | 0.017 | 5.94 (3.28–10.7) | <0.001 |

Complete-case sample used for all models (n = 1,619). The reference outcome is no headache-relevant medication.
RRR, relative risk ratio from multinomial logistic regression; CI, confidence interval; HIT-6, Headache Impact Test-6; mTBI, mild traumatic brain injury; PCL-5, PTSD Checklist for DSM-5; PHQ-9, Patient Health Questionnaire-9; PTSD, posttraumatic stress disorder; TBI-QOL, Traumatic Brain Injury–Quality of Life.

**Table S6.** Fully adjusted multinomial logistic regression of headache medication pattern (Model 3; n = 1,619)

| Characteristic | Acute only | | Preventive only | | Combination therapy | |
| --- | --- | --- | --- | --- | --- | --- |
|  | RRR (95% CI) | p-value | RRR (95% CI) | p-value | RRR (95% CI) | p-value |
| Lifetime mTBI |  |  |  |  |  |  |
| No mTBI | 1.00 (Ref) |  | 1.00 (Ref) |  | 1.00 (Ref) |  |
| 1 mTBI | 1.14 (0.74–1.74) | 0.6 | 1.43 (0.73–2.78) | 0.3 | 0.91 (0.54–1.56) | 0.7 |
| 2+ mTBIs | 1.19 (0.81–1.74) | 0.4 | 1.61 (0.87–2.95) | 0.13 | 1.05 (0.66–1.69) | 0.8 |
| Age, years | 1.04 (1.03–1.05) | <0.001 | 1.03 (1.01–1.05) | <0.001 | 1.05 (1.03–1.07) | <0.001 |
| Sex |  |  |  |  |  |  |
| Female | 1.00 (Ref) |  | 1.00 (Ref) |  | 1.00 (Ref) |  |
| Male | 0.82 (0.57–1.17) | 0.3 | 0.63 (0.39–1.03) | 0.068 | 0.63 (0.42–0.95) | 0.028 |
| Hypertension |  |  |  |  |  |  |
| No | 1.00 (Ref) |  | 1.00 (Ref) |  | 1.00 (Ref) |  |
| Yes | 1.12 (0.86–1.46) | 0.4 | 2.72 (1.90–3.90) | <0.001 | 2.96 (2.18–4.03) | <0.001 |
| PTSD (PCL-5) |  |  |  |  |  |  |
| No PTSD (<33) | 1.00 (Ref) |  | 1.00 (Ref) |  | 1.00 (Ref) |  |
| PTSD (≥33) | 0.72 (0.51–1.02) | 0.065 | 1.16 (0.71–1.89) | 0.6 | 0.90 (0.60–1.36) | 0.6 |
| Depression (PHQ-9) |  |  |  |  |  |  |
| No depression | 1.00 (Ref) |  | 1.00 (Ref) |  | 1.00 (Ref) |  |
| Mild | 0.91 (0.63–1.30) | 0.6 | 0.83 (0.48–1.42) | 0.5 | 0.85 (0.53–1.35) | 0.5 |
| Moderate | 1.02 (0.67–1.57) | >0.9 | 1.01 (0.54–1.90) | >0.9 | 0.78 (0.45–1.35) | 0.4 |
| Moderate/Severe | 0.94 (0.54–1.63) | 0.8 | 1.20 (0.55–2.59) | 0.6 | 1.01 (0.53–1.96) | >0.9 |
| TBI-QOL Anxiety, T-score | 0.99 (0.97–1.00) | 0.15 | 0.98 (0.95–1.00) | 0.078 | 0.98 (0.96–1.00) | 0.10 |
| Sleep quality |  |  |  |  |  |  |
| Good sleep | 1.00 (Ref) |  | 1.00 (Ref) |  | 1.00 (Ref) |  |
| Poor sleep | 1.33 (0.87–2.05) | 0.2 | 1.85 (0.90–3.78) | 0.093 | 1.77 (0.94–3.34) | 0.078 |
| Headache impact (HIT-6) |  |  |  |  |  |  |
| Little to no impact | 1.00 (Ref) |  | 1.00 (Ref) |  | 1.00 (Ref) |  |
| Some impact | 1.09 (0.72–1.65) | 0.7 | 1.09 (0.58–2.03) | 0.8 | 1.72 (0.91–3.25) | 0.10 |
| Substantial impact | 1.22 (0.77–1.93) | 0.4 | 0.97 (0.47–1.99) | >0.9 | 1.93 (0.97–3.83) | 0.062 |
| Severe impact | 1.71 (1.14–2.58) | 0.010 | 1.77 (0.96–3.25) | 0.067 | 4.33 (2.36–7.95) | <0.001 |
| TBI-QOL Pain interference, T-score | 1.05 (1.02–1.07) | <0.001 | 1.03 (1.00–1.06) | 0.069 | 1.06 (1.03–1.09) | <0.001 |

Complete-case sample used for all models (n = 1,619). The model includes all variables shown. The reference outcome is no headache-relevant medication.
RRR, relative risk ratio from multinomial logistic regression; CI, confidence interval; HIT-6, Headache Impact Test-6; mTBI, mild traumatic brain injury; PCL-5, PTSD Checklist for DSM-5; PHQ-9, Patient Health Questionnaire-9; PTSD, posttraumatic stress disorder; TBI-QOL, Traumatic Brain Injury–Quality of Life.

**Table S7.** Sensitivity analyses: association of lifetime mTBI with headache medication use under alternative medication classifications and among participants with a recorded headache diagnosis

| Model | Lifetime mTBI (vs. none) | Acute only vs. none | Preventive only vs. none | Combination therapy vs. none |
| --- | --- | --- | --- | --- |
| *A. Restrictive medication classification (n = 1,619): relative risk ratio (95% CI)* | | | | |
| Model 1 | 1 mTBI | 1.20 (0.81–1.79) | 1.80 (0.83–3.89) | 1.33 (0.75–2.36) |
|  | 2+ mTBIs | 1.29 (0.91–1.82) | 1.82 (0.90–3.69) | 1.69 (1.02–2.81)* |
| Model 3 | 1 mTBI | 1.06 (0.71–1.60) | 1.45 (0.66–3.19) | 0.96 (0.53–1.76) |
|  | 2+ mTBIs | 1.07 (0.74–1.53) | 1.39 (0.67–2.87) | 1.13 (0.66–1.93) |
| *B. Any relatively headache-specific treatment vs. none (n = 1,619): logistic regression, odds ratio (95% CI)* | | | | |
| Model 1 | 1 mTBI | OR 1.36 (0.81–2.31), any headache-specific treatment vs. none | | |
|  | 2+ mTBIs | OR 1.47 (0.93–2.39), any headache-specific treatment vs. none | | |
| *C. Participants with a recorded headache diagnosis in VA health records (n = 757): relative risk ratio (95% CI)* | | | | |
| Model 1 | 1 mTBI | 1.22 (0.64–2.31) | 2.34 (0.79–6.92) | 1.17 (0.58–2.37) |
|  | 2+ mTBIs | 1.14 (0.65–2.02) | 2.67 (0.98–7.27) | 1.36 (0.73–2.53) |
| Model 3 | 1 mTBI | 1.23 (0.64–2.36) | 2.15 (0.71–6.47) | 0.98 (0.47–2.05) |
|  | 2+ mTBIs | 1.24 (0.69–2.24) | 2.46 (0.89–6.82) | 1.15 (0.60–2.22) |

Model 1: adjusted for age and sex. Model 3: Model 1 plus hypertension, PTSD, depression, anxiety, sleep quality, headache impact, and pain interference. * p < 0.05.
**A.** Multinomial logistic regression with the outcome redefined under the restrictive classification (Supplementary Table S2).

**B.** Logistic regression of any triptan, gepant/ditan, ergot derivative, CGRP monoclonal antibody/atogepant, or botulinum toxin use vs. none of these.

**C.** Multinomial logistic regression using the primary medication classification, restricted to participants with headache diagnosis (migraine, tension-type, post-traumatic, or unspecified headache) in VA health records (available for 1,150 participants); CI, confidence interval; mTBI, mild traumatic brain injury

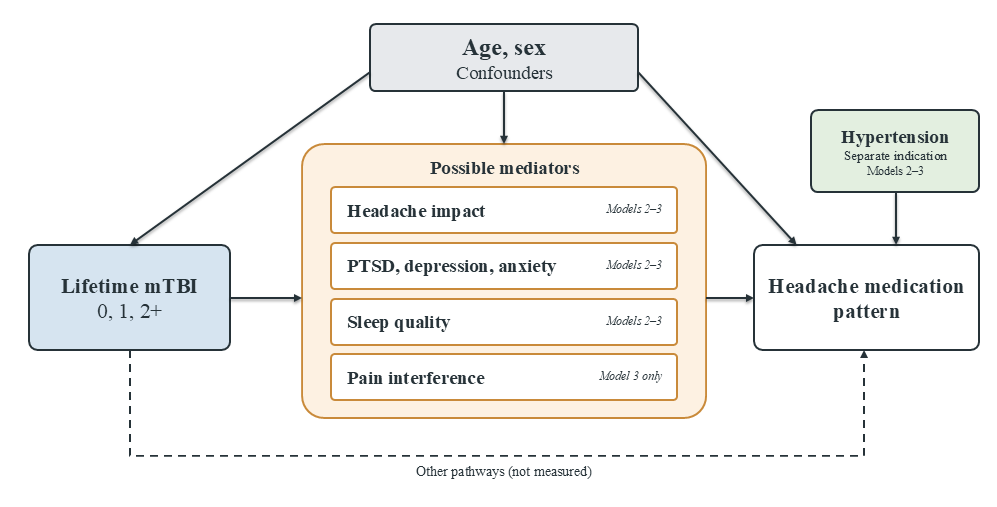

**Figure S1.** Conceptual diagram of hypothesized relationships between lifetime mTBI and headache medication pattern.

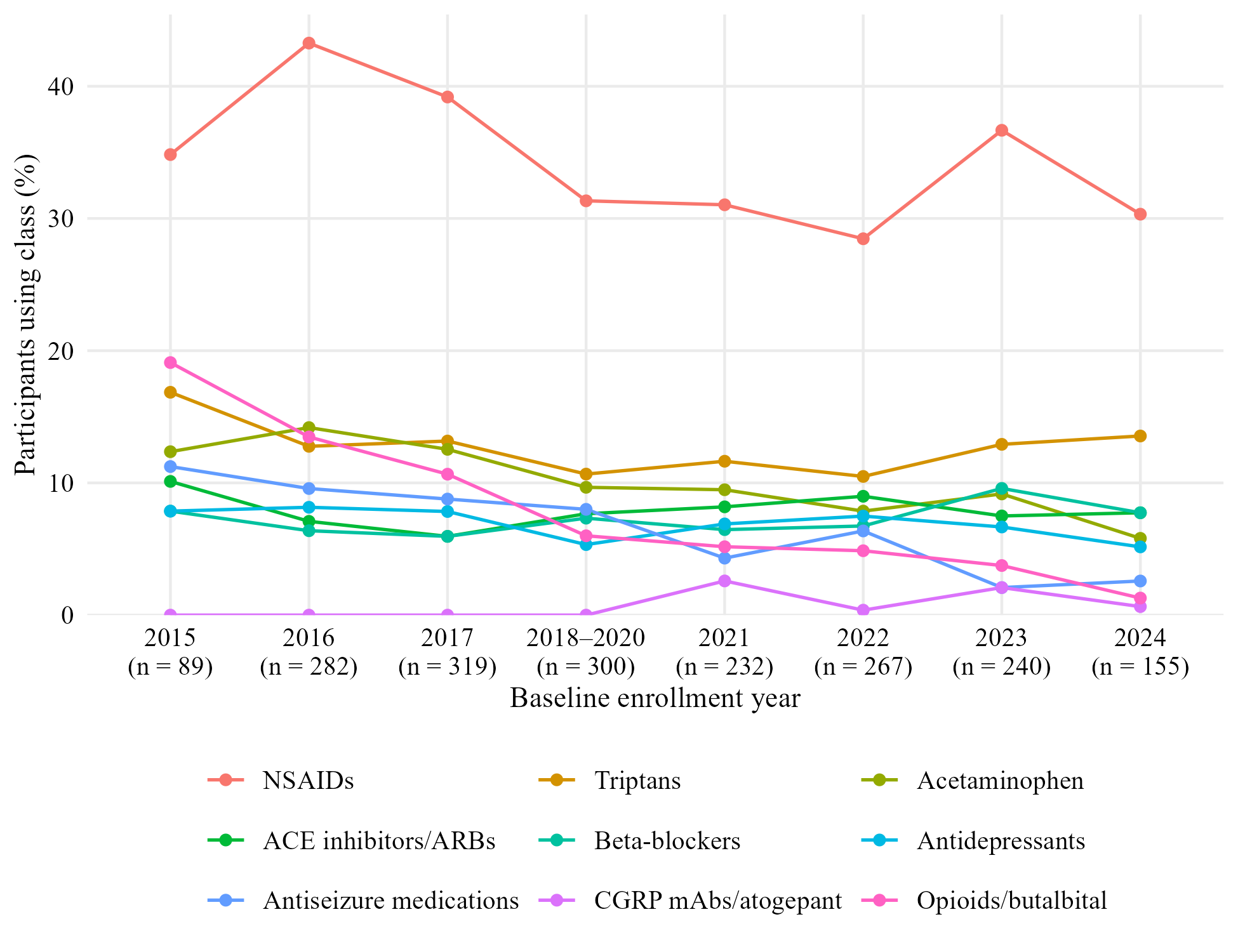

**Figure S2**. Use of headache-relevant medication classes by baseline enrollment year (N = 1,884).

ACE, angiotensin-converting enzyme; ARBs, angiotensin II receptor blockers; CGRP mAbs, calcitonin gene-related peptide monoclonal antibodies; NSAIDs, nonsteroidal anti-inflammatory drugs.
